# Design and Usability Evaluation of a Digital Guideline Management Application for a Pediatric Cardiac Center

**DOI:** 10.64898/2026.05.24.26353982

**Authors:** Brianna Heidenreich

## Abstract

**Background:** Complex cases in specialized pediatric care require consistent adherence to evidence-based clinical pathways and protocols to ensure safe, high-quality, and equitable care. Currently, clinical pathways and supporting documentation are frequently distributed across multiple platforms, leading to fragmentation. Human-centered design principles can guide the development of healthcare technologies that minimize cognitive load and support rapid, efficient access to relevant information in clinical settings. The purpose of this study is to design and evaluate perceived usability of a pediatric cardiac center digital guideline management system that is embedded within the electronic health record leveraging human-centered design.

**Methods:** This study used a mixed-methods usability evaluation to assess a digital guideline management system prototype embedded into clinical workflow. Through human-centered design principles, the prototype provides a centralized digital document library that organizes cardiac-specific clinical pathways, guidelines, procedures, and related resources. A small but diverse sample, encompassing a wide variety of roles and clinical areas within the pediatric cardiac center, was recruited to evaluate the perceived usability of the prototype. Usability was evaluated by stakeholders using the validated System Usability Scale (SUS) with additional optional questions to understand perceptions of the information architecture and clinical value.

**Results:** Preliminary usability testing showed a mean SUS composite score of 76.5, indicating above average usability. Questions related to the complexity of the system and user confidence received high scores across participants. Lower scores were observed for questions related to usage frequency and ability to learn the system very quickly.

**Conclusion:** Leveraging human-centered design when building a digital guideline management system embedded within clinical workflow revealed positive perception from participants. By centralizing access to clinical resources, this prototype can reduce current-state fragmentation. Further evaluation of larger samples is needed to develop a list of future recommendations.

## INTRODUCTION

Pediatric congenital and acquired heart disease represents a significant public health burden, with an estimate of one child born with a congenital heart defect every 15 minutes in the United States. Approximately 25% of these children have a critical heart defect, requiring serious interventions within the first year of life^1^. Advances in pediatric cardiology and cardiac surgery have improved survival, resulting in increasingly complex patient populations managed across inpatient, intensive care, perioperative, and outpatient settings. This complexity in specialized pediatric care requires consistent adherence to evidence-based clinical pathways and protocols to ensure safe, high-quality, and equitable care ^2,3,4,5,6^.

Despite the availability of guidelines, pediatric cardiac care teams often experience difficulty accessing the most relevant and up-to-date guidance at the point of care ^10, 27, 28^. Clinical pathways and supporting documentation are frequently distributed across multiple platforms, including shared document repositories, locally stored documents, and external journal publications, leading to fragmentation^7^. Information that is challenging to access is less likely to be used in clinical practice. Studies have shown that physicians seek answers to approximately 50% of the clinical questions that arise during patient care; However, only a subset of these questions is resolved^17^. Numerous barriers may be faced when seeking information to help guide patient care, including time limitations and uncertain knowledge accuracy ^5, 18, 19^. In high-acuity environments, fragmentation can lead to increased cognitive burden, patient safety risks, and variation in care delivery^8, 9, 10^.

From a population health perspective, inefficient access to appropriate clinical knowledge represents a systems-level issue that impacts equitable outcomes, preventable adverse events, clinician workload, and resource inefficiencies across patient populations ^8, 12^. EHR-integrated digital tools have the potential to improve accessibility of relevant clinical guidelines and improve patient outcomes when considering the end user experience. Human-centered design principles emphasize user-informed design methodology that identify risks early on and aim to improve workflow efficiency ^9, 21,22^. When designed thoughtfully, clinical decision support and digital guideline management systems have demonstrated improvement in the standardization of care, clinician efficiency, and patient safety ^20.^ When users are not considered in the early stages of technological development, design interfaces may contribute to documentation errors, documentation burden, and alert fatigue, hindering the adoption of the system ^16, 21, 23, 24^.

The purpose of this study was to design and evaluate the usability of a human-centered digital guideline management prototype that supports pediatric cardiologists and associated care team members in delivering safe, evidence-based, and standardized care at the bedside. This paper addresses an informatics challenge that focuses on patient safety, clinical experience, and healthcare system efficiency. By reducing fragmentation and improving access to institutional clinical guidelines, the proposed digital guideline management system may reduce clinical cognitive burden, improve workflow efficiency, promote consistent use of institutional best practices, and support safer and more standardized care delivery for medically complex pediatric cardiac populations.

## METHODS

### Study Design

This study used a mixed-methods usability evaluation to assess a digital guideline management prototype designed to support clinician access to institutional clinical resources. The prototype was developed using an existing EHR dashboard infrastructure to simulate a centralized, point-of-care digital library. The dashboard organized clinical pathways, guidelines, procedures, and supporting resources in a structured manner that incorporated navigation-focused design elements to support strong usability. The study focused on evaluating perceived usability and workflow alignment. A human-centered design approach guided both the development and evaluation of the prototype, prioritizing the demands within clinical workflow and ease of use.

### Data Sources

A survey instrument was developed and administered using Research Electronic Data Capture (REDCap). The survey incorporated System Usability Scale (SUS) evaluation, a validated 10-item questionnaire using a 5-point Likert scale, to assess perceived usability of the prototype. To interpret SUS scores, adjective-based groupings were applied: < 51.6 indicating poor, 51.7-71 signifying an okay score, 71.1-77.1 representing a good (above average) score, 77.2-84 describing an excellent score, and > 84 demonstrating best imaginable score ^25, 26^. Additional optional binary-response questions were included to capture feedback on design and information architecture. Participants were expected to access the prototype directly within the EHR environment and complete the survey to provide feedback on the usability, organizational structure, and navigation features of the prototype.

### Study Population

Twenty clinical and operational stakeholders from a pediatric cardiac center at a large academic children’s hospital in the United States were asked to participate in evaluating the digital guideline management application prototype. Out of the 20 stakeholders, 5 completed the survey. Participants were purposively selected to represent a range of roles and care settings within the institution, including physicians, advanced practice providers, nursing staff, and operational leaders across the inpatient, outpatient, and procedural care environments. The sample size was consistent with prior usability literature that suggested that a relatively small sample population can identify a substantial proportion of usability issues ^22, 31^.

### Prototype Development

Human-centered design principles drove the development of the prototype, targeting intuitive navigation, efficient informational retrieval, and workflow alignment. The design of the first prototype prioritized mechanisms to reduce the number of clicks required to access relevant resources, improving the organization of institutional-specific content and providing an efficient information retrieval environment (Figure 1) ^6, 18, 23, 29, 30^. The prototype was organized into three columns, with each sub header color coded with the corresponding column’s title. Dropdown menus were provided to organize the documents by service area and topics. Selection of the dropdown menu displayed relevant hyperlinks to institutional documents. Hover-over synopsis functionality provided a brief description of the linked resource to support the identification of document relevance.

**Figure 1.**
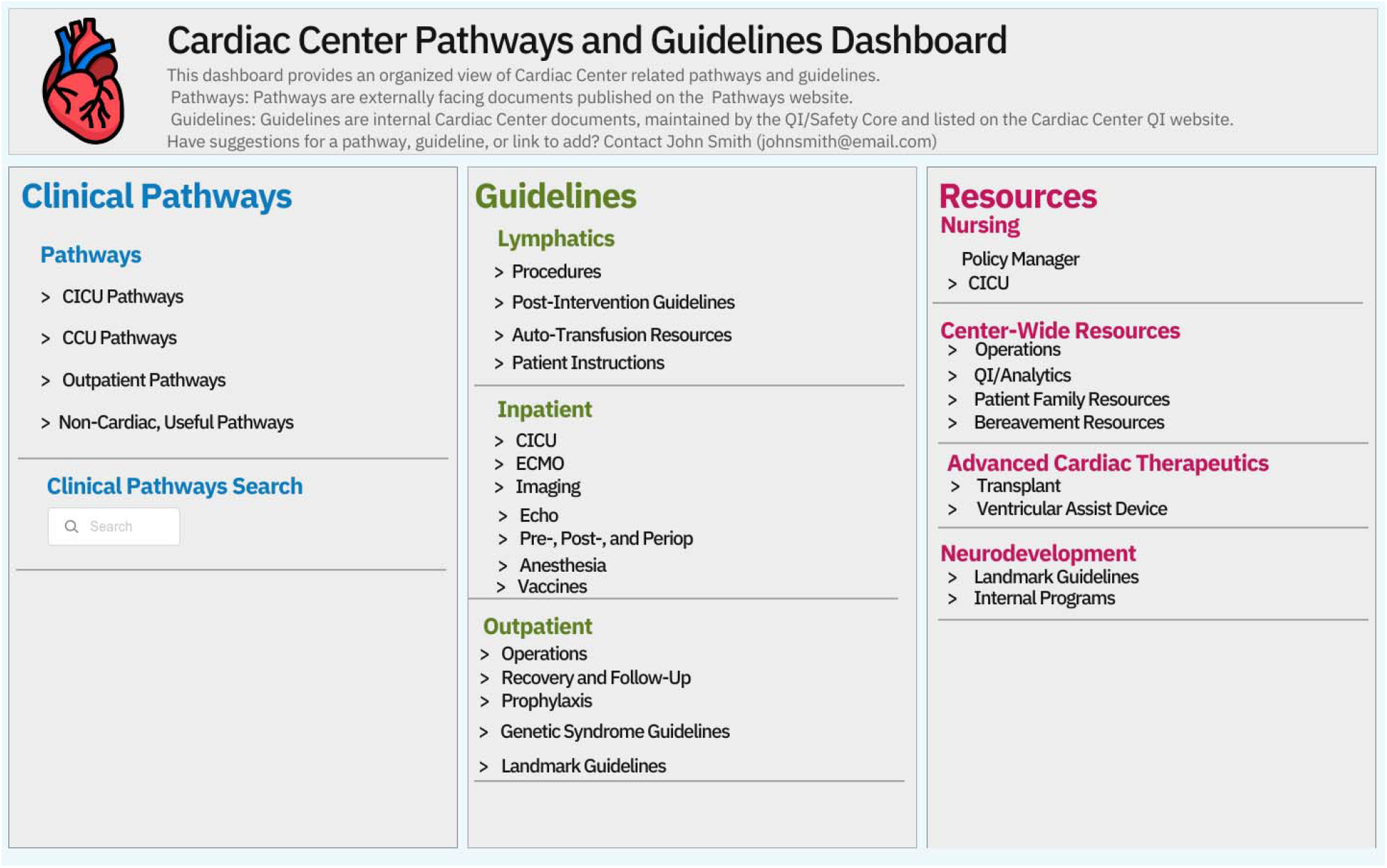
First Iteration of Prototype. The prototype organized institutional resources into three primary categories: Clinical Pathways, Guidelines, and Resources. Dropdown navigation menus, indicated by the “>“ symbol, were used to categorize documents by clinical area and topic for ease of navigation.

### Variables

The primary outcome was perceived system usability, measured by SUS scores. Responses were converted into composite SUS scores ranging from 0-100. Scores were interpreted using established benchmarks, with overall scores above 77.2 indicating excellent usability. Secondary variables included participant responses to individual SUS items to assist in identifying the strengths and opportunities of improvement in a future iteration of the prototype. Additional variables stemmed from study-specific questions to assess design and information architecture to further evaluate the perception of usability when seeking clinical information. Participant role was collected as a categorical variable to describe the study sample.

### Statistical Analysis

Given the exploratory nature of the study, descriptive statistics were used for the analysis of the prototype. Individual SUS-item responses were reviewed to identify patterns across core usability themes, including perceived ease of use, system complexity, and user confidence. Once composite SUS scores were calculated, summary statistics (mean, standard deviation, median, and range) were calculated to describe overall perceived usability of the prototype. Binary-response variables to assess design and information architecture of the prototype were analyzed through proportions to identify the percentage of participants reporting positive perceptions of the prototype.

### Ethical Considerations

A Human Subjects Research (HSR) determination form was submitted through the City University of New York School of Public Health and Health Policy Human Research Protection Program (HRPP). Based on institutional review, the project was determined not to constitute Human Subjects Research, and Institutional Review Board (IRB) approval was not required. Any information related to the participants has been de-identified.

## RESULTS

### Participant Characteristics

A total of 5 participants completed usability testing of the proposed solution, representing a multidisciplinary sample to properly capture the needs across the Cardiac Center. Participants included physicians (20%), registered nurses (40%), and stakeholders whose primary purpose is centered around quality improvement (QI), operations, and data analytics (40%) (Table 1).

**Table 1.** Participant Characteristics.

| Role | Participants (N = 5) |
| --- | --- |
| Physician | 20% (n = 1) |
| Advanced Practice Provider | 0% (n =0) |
| Registered Nurse | 40% (n = 2) |
| QI/ Operations/ Data Analytics | 40% (n =2) |

QI: Quality Improvement. Participant roles represent multidisciplinary stakeholders within the pediatric cardiac center that may interact with the resource who completed the usability evaluation of the digital guideline management prototype. Percentages were calculated from the total number of study participants (N =5).

### Usability Outcomes

The prototype demonstrated positive perceived usability with a mean SUS score of 76.5 (SD = 4.2), with scores ranging from 72.5-82.5. The higher the overall composite score, the greater that the perceived usability of the system would be. This score can be condensed into categories to qualitatively describe the meaning of the numerical value. Based on established benchmarks, this score corresponds to good (above average) usability (Table 2).

**Table 2.** SUS Summary Statistics.

| Metric | Value |
| --- | --- |
| Mean SUS Score | 76.5 |
| Standard Deviation | 4.2 |
| Median | 77.5 |
| Range | 72.5-82.5 |
| Interpretation | Good (Above average) |

SUS: System Usability Scale. Composite SUS scores were calculated using standard SUS scoring methodology, creating a possible range of scores from 0 to 100, with higher scores indicating higher
perceived usability. Interpretation categories were based on established SUS adjective rating scales, with scores ranging from 71.1 to 77.1 representing good, above average perceived usability.

The distribution of the composite SUS across participants is shown in Figure 2. Scores showed minimal variation between participants, indicating a similar perception of prototype system usability amongst the entire sample population (Figure 2). Questions related to the complexity of the system and user confidence received consistently high scores across participants. Lower scores were observed for questions related to usage frequency and ability to learn the system very quickly, impacting the overall score in usability evaluation.

**Figure 2.**
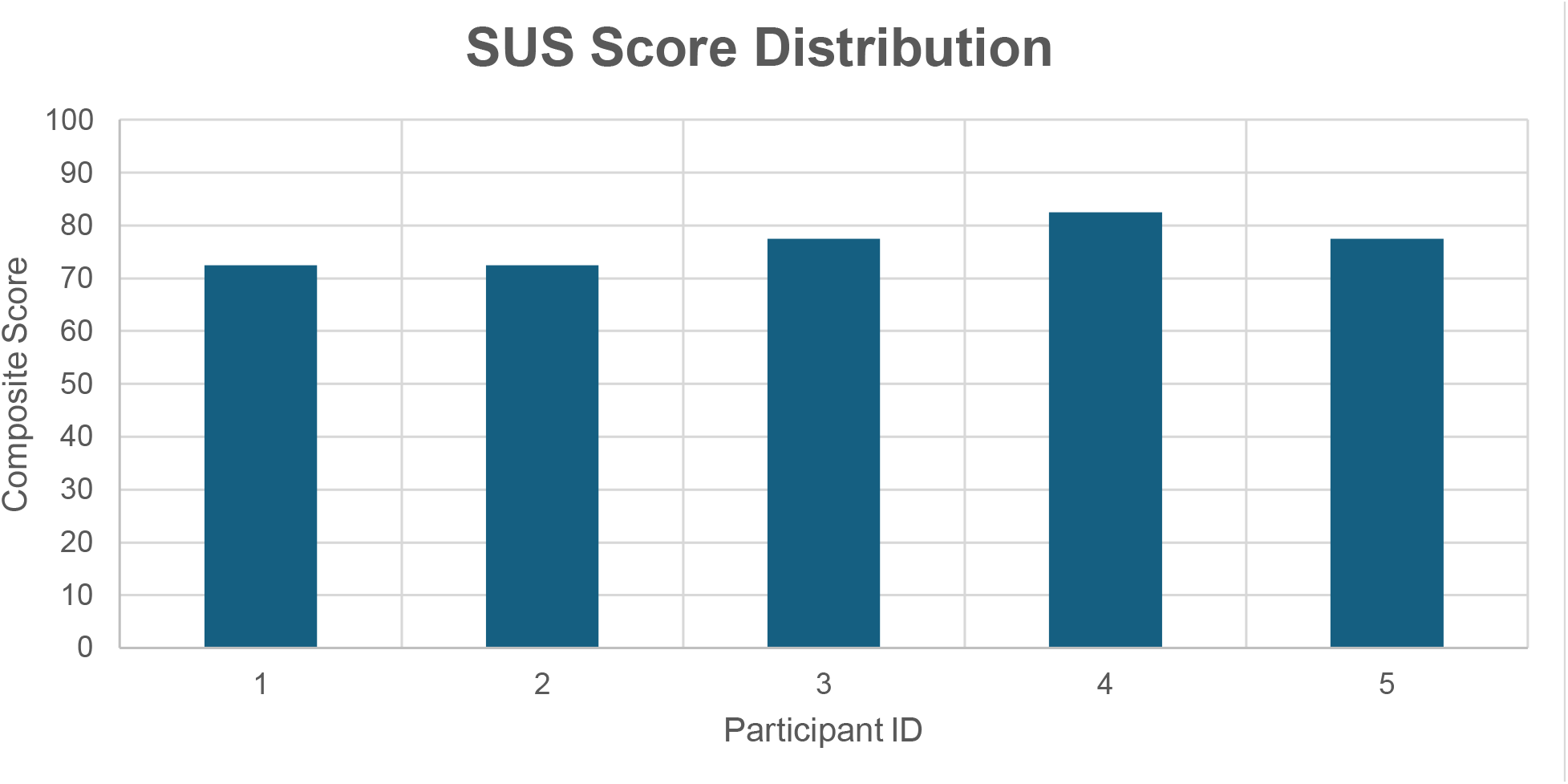
SUS Score Distribution Each bar represents individual participant composite SUS scores (0-100 scale), with higher scores indicating higher perceived usability of the digital guideline management systems.

### Design and Information Architecture

Participants reported predominantly positive feedback with the information architecture of the prototype. There was a mixed perspective regarding the categories that were selected to index the digital library. Column headers and sub headers were positively perceived as they provided clear and meaningful context. Users felt that the dropdown feature embedded in the sub headers was an effective way to organize the documents. The hover-over synopsis feature, which provides a brief highlight of document purpose, was viewed as a helpful addition to the resource. There was a reported perception of indifference towards the overall adoption of the digital guideline management system as participants felt neutral on the resource (Table 3).

**Table 3.** Design and Information Architecture Breakdown.

| Domain | Item | Response Type | % Positive Response |
| --- | --- | --- | --- |
| Information Architecture | Categories are intuitive and appropriate | Yes/No | 80% (4 yes, 1 no) |
| Information Architecture | Headers and sub headers are clear and meaningful | Yes/No | 100% Yes |
| Navigation | Dropdowns effectively organize documents | Yes/No | 100% Yes |
| Usability Enhancement | Hover-over synopsis is helpful | Yes/No | 100% Yes |
| Adoption | Would use system frequently | Likert (Agree/ Strongly Agree) | 100% Neutral |

Responses were collected through supplemental usability survey questions administered after prototype review. Positive responses were defined as “yes” in binary responses and “Agree” or “Strongly Agree” for Likert-scale items.

## DISCUSSION

This study aimed to evaluate the perceived usability of a digital guideline management system prototype leveraging human-centered design principles. Findings demonstrated above-average usability, with a mean SUS score of 76.5 and a low standard deviation (4.2), indicating low variability across the entire sample population. Participants also provided supplemental feedback on the information and design architecture of the prototype. Participants indicated that key design features, including navigation structure and document organization, were clear, meaningful, and helpful. Despite the positive perception towards the design and usability of this digital resource, the entire sample population reported neutral perceptions regarding the frequency of system usage. This further suggests that the general design may support the needs of the user, but additional refinement and real-world integration may be necessary to better understand potential long-term adoption.

These findings strengthen what existing literature has shown by stressing the importance of usability testing and workflow alignment before investing in the adoption of digital tools. Previous studies have demonstrated that fragmented access to clinical practice guidelines impacts clinical cognitive burden and workflow inefficiencies. Clinical pathways, guidelines, and other relevant resources are available across multiple platforms, creating unnecessary barriers to efficient information retrieval of appropriate clinical information during patient care. This study sought to address these barriers and support more efficient access to evidence-based best practice resources.

There are numerous strengths to this study. Firstly, usability testing was conducted within an existing dashboard infrastructure that is embedded in the EHR, allowing participants to contextualize the prototype directly in the clinical environment as opposed to an external platform. The study also recruited participants that represented diverse clinical settings and roles within the pediatric cardiac center, allowing for a diverse set of end users with unique clinical perspectives to provide feedback. Lastly, the study utilized quantitative measures through a validated scoring system to evaluate the usability and user experience of the proposed solution, with supplemental qualitative feedback to provide broader perspective on the perceptions of the prototype.

Although existing literature has shown that a relatively small number of participants can identify a substantial proportion of usability issues, this sample size was limited to a single pediatric cardiac center and may reduce the applicability of the results to other settings and institutions. The study evaluated a prototype rather than a fully implemented workflow-integrated solution. Because of this, the findings focused on perceived usability and anticipated workflow integration, rather than a direct measure of long-term adoption and workflow efficiency.

From a population health perspective, improving access to evidence-based clinical guidelines may improve efficiency of clinical information retrieval and decrease fragmentation across an entire clinical system. Clinicians in specialized care environments, such as pediatric cardiac centers, often rely on internal pathways and guidelines to guide complex clinical care. Poorly integrated digital systems and fragmented information environments may contribute to clinician cognitive burden, workflow disruption, excessive screen navigation, and documentation burden. All of these clinical experiences are associated with EHR-related fatigue and reduced efficiency within high-acuity clinical environments that may negatively impact across subsequent patient encounters^13,14,15,16^. Because of this, effective information access systems are important in specialized care environments as they support safe, high-quality, and equitable care.

Future studies should expand upon usability testing to a larger sample population, including broader multidisciplinary representation across specialties and institutions. Additional studies will need to focus on the evaluation of centralized digital guideline management systems in real-world clinical practice to evaluate whether they improve information retrieval efficiency and user experience over time. Future iterations of the prototype can explore alternative platforms that have the capability to incorporate more advanced design functions, such as enhanced search functionalities, clinical scenario customization, and bedside accessibility, to determine the most appropriate long-term platform for workflow integration and deployment. Future development of digital guideline management systems should prioritize not only evidence-based resources, but also how they can be properly integrated into the clinical workflow.

## CONCLUSION

Leveraging human-centered design when building a digital guideline management system embedded within clinical workflow demonstrated strong usability and a highly favorable reception among clinical stakeholders. By centralizing access to clinical resources, this prototype offers a viable strategy to mitigate systemic information fragmentation. However, further evaluation of larger samples and alternative technical architectures is needed prior to widespread adoption within live clinical workflows.

## Data Availability

All data produced in the present study are available upon reasonable request to the authors

## ACKNOWLEDGMENTS

The author would like to acknowledge Susan Ferry MHL, RRT-NPS; Amy Schiltz MD, MSCE; Thomas Dietzman MD; Michael Goldsmith MD; and David Hehir MD, MS for their guidance, mentorship, and continued support throughout development of this project. The author would also like to acknowledgethe multidisciplinary stakeholders across the pediatric cardiac center who participated in usability evaluation and provided valuable clinical and operational perspectives that informed prototype refinement.

## CONFLICT OF INTEREST STATEMENT

The author declares no conflicts of interest.

## FUNDING STATEMENT

No external funding was received for this study.

## Notes

### Competing Interest Statement

The authors have declared no competing interest.

### Author Declarations

A Human Subjects Research (HSR) determination form was submitted through the City University of New York School of Public Health and Health Policy Human Research Protection Program (HRPP). Based on institutional review, the project was determined not to constitute Human Subjects Research, and Institutional Review Board (IRB) approval was not required.

